# Evaluation of the disinfecting capacity of ozone in emergency vehicles

**DOI:** 10.1101/2020.05.24.20111666

**Authors:** Jorge Biurrun Cía, Begoña García Martínez, Andrea Perez Montero, Grazyna Kochan, David Escors, José Crespo Martinez, Iñigo Lasa Uzcudun, Alfredo Echarri Sucunza

## Abstract

**Objective:** As a consequence of the health crisis arising from the SARS-CoV-2 coronavirus pandemic, ozone treatments are being applied as disinfectant in emergency vehicles, without objective evidence on its efficacy. Here we evaluate the efficacy of ozone treatment over bacterial strains and virus-like particles.

**Method:** A preparation of a lentiviral vector (lentivector) and dried cultures of two bacterial strains (gram + *Staphylococcus aureus* and gram - *Salmonella enterica ser*. Enteritidis) were placed inside an ambulance at two different locations. The interior of the vehicle was subjected to 10 min and 20 min treatments (3 and 6 times the recommended time by the manufacturer). Following the treatments, lentivector preparations were titrated, and viable bacteria (colony forming units, CFUs) counted and compared to pre-treatment titers and infectious CFUs of the same lysates and cultures.

**Results:** None of the treatments significantly reduced either lentivector titer or the number of viable bacteria.

**Conclusions:** At least in the analyzed conditions and for the microorganisms used in this study, it can be concluded that ozone treatment is not advisable for the disinfection of emergency vehicles.

## INTRODUCCIÓN

As a consequence of the pandemic caused by SARS-CoV-2 the disinfection of emergency vehicles has become a priority, as this is the main transport means of infected patients. Therefore, these vehicles can be a potential source for new infections of patients not related to SARS-CoV-2 infections.

The different governmental organizations, emergency services and private transport companies of biomedical materials are putting in place several measures for disinfecting vehicles. Ozone cannons are currently the most used method for disinfecting the surfaces of vehicles, due to their rapid and straightforward application. Ozone is an oxidizing gas with a demonstrated disinfecting capacity in aqueous solutions, and it is widely used for disinfecting food and water (1-4). However, its efficacy to disinfect surfaces by nebulization has not been properly tested. Therefore, the Spanish Ministery of Health does not include ozone in the list of authorized virucides for disinfecting surfaces https://www.mscbs.gob.es/profesionales/saludPublica/ccayes/alertasActual/nCov-China/documentos/Listado_virucidas.pdf).

Here we have tested the efficacy of ozone disinfection using ozone cannons as currently applied in ambulances. To this end, we have studied its efficacy in reducing the viability of three model microorganisms: a lentivector and two bacterial pathogens *(Staphylococcus aureus* and *Salmonella entérica)*. Lentivectors are virus-like particles (VLPs) that can be used as a biosafe model to evaluate disinfectant over viral infectious agents. Coronaviruses contain a single-stranded RNA genome packaged by the nucleoprotein, which is in turn enclosed in a membrane envelope containing the S, M and E proteins (5). HIV-1-derived lentivectors show a similar structure to SARS-CoV-2, containing a transfer single-stranded RNA packaged by the nucleocapsid, capsid and matrix proteins, enclosed in a membrane envelope containing the vesicular stomatitis virus G protein (VSV-G) (6). Lentivectors can be used as transfer vehicles of reporter genes such as GFP into susceptible cells, which allows the quantification of infectious viral particles by evaluating GFP expression. Lentivectors cannot propagate in cell cultures, so they represent a biosafe model agent for RNA viruses such as SARS.

In addition, these treatments have also been proposed to as broad spectrum disinfectants. Hence, our study has also tested the capacities of ozone treatments over two model bacterial pathogens, *Staphylococcus aureus* y *Salmonella enterica*. These gram-positive and gram-negative strains are recomended by American (EPA) and European (EN) norms (7) (8) for the evaluation of the efficacy of disinfecting treatments over surfaces.

## METHODS

The pSIN-GFP lentivector was prepared by the three-plasmid co-transfection method as previously described. This lentivector encodes the GFP gene under the control of the SFFV promoter (9). Lentivector preparations were titrated in 293T cell cultures as described by Selden *et al*. (10). *Staphylococcus aureus* strain MW2 *(S. aureus* MW2) (11) and *Salmonella enterica ser*. Enteritidis strain 3934 *(S*. Enteritidis 3934) (12) were grown overnight in TSB and LB medium at 37°C in shakers (200 rpm), respectively. Then, 1 ml of cultures were retrieved by centrifugation, washed in PBS thrice and resuspended in 1 ml of PBS. 25 μl droplets of bacteria suspensions were air-dried in Petri dishes under sterile conditions.

A medicalized ambulance of 10,79 m^3^ was used for testing ozone treatments. Ozone was produced with an ozone cannon (Industrial Global Supply S.L.) with a production capacity of 16000 mg/h. The ozone cannon was placed on the floor, and the production cannon oriented towards the stretcher (Figure 1). Open petri dishes containing 1 ml of the pSIN-GFP lentivector stock in DMEM and containing dried bacterial cultures were placed on the stretcher at two different locations. A “proximal” location next to the ozone cannon, and a “distal” location from the ozone cannot (Figure 1). Ozone levels inside the ambulance were monitored using a Dräger Pac 8000. The ambient humidity was also measured with a hygrometer (ThermoPro) (Figure 1). Two different treatments were carried out, differing in duration (10 and 20 min), following the specifications provided by the manufacturer of the ozone cannon, by calculating the exposure time as a function of the target volume (0,3 min/m^3^ in emergency vehicles). 3,23 min would suffice for the complete disinfection of the ambulance used for this study according to these specifications.

**Figure 1.**
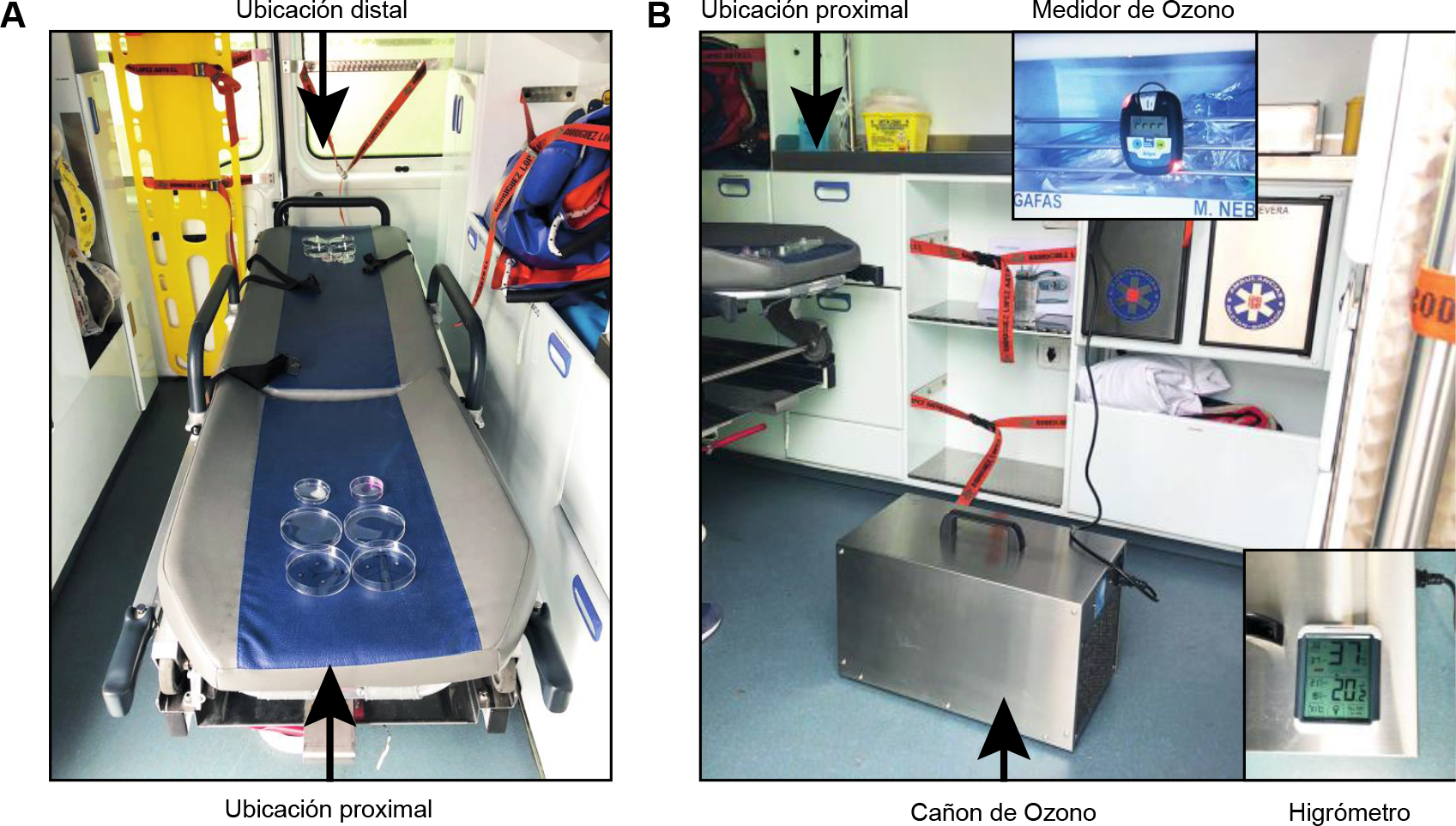
Environmental context and experimental setup. ***(A)*** *Spacial location of the lentivector stocks and bacterial cultures on the stretcher within the ambulance, at the two sites (proximal and distal) referenced to the position of the ozone cannon*. ***(B)*** *Position of the ozone cannon within the ambulance. The ozometer was placed in the upper shelves as shown in the picture. The picture shows the moment in which the ozometer shows ozone levels superior than 10 parts per million (ppm), indicating the saturation of the ozometer and the warning lights. The hygrometer was introduced in the ambulance before and after the tests to register the humidity range during the experiments*.

Accordingly, 10 and 20-min treatments correspond to a 3 and 6-fold increase in the recomended treatment time, respectively.

The lentivector stocks were immediately titrated following the treatments by transducing cultures of 293T cells (5×10^5^ cells). Three days later, the number of lentivector infectious units were calculated by examining the proportion of GFP positive cells by fluorescence microscopy and flow cytometry, following published procedures (10); (9). Mean and standard deviations were calculated from four independent quantifications for each experiment at the two different locations. Data was compared to the same untreated lentivector stocks.

Bacterial cultures were rehydrated after the treatments in 100 μl PBS and the number of viable cells was calculated and compared to untreated bacterial samples. Quantifications were carried out by 10-fold serial dilutions of each simple, and performing cell counts in triplicates. Means and standard deviations were calculated from three independent bacterial cultures.

As a control, UV-C light was used as a disinfectant agent, which is demonstrated to be efficacious for disinfecting surfaces (13-16). Hence, open Petri dishes containing 1 ml of pSIN-GFP lentivector stock in DMEM as well as Petri dishes with desiccated bacterial cultures were located in the interior of a laminar flow cell culture cabinet and subjected to UV-C irradiation for 20 min (TUV 30W, 100μW/cm^2^ at 1 m). Lentivector titres and bacterial counts were performed as described above.

## RESULTS

During the test, ozone levels higher than 10 ppm were reached within the ambulance for both 10 and 20-min treatments (Figure 1). The humidity during the experiments ranged from 37 to 48% (Figure 1). Ozone treatments barely affected the lentivector titres (Figure 2 and 3). Independently of the time and location from the ozone cannon, the differences were always inferior to a logarithm (10^-1^) (Figure 3). In contrast, the UV light treatment reduced the lentiector titre below the detection limit of the titration technique (< 5000 partículas/ml), reaching at least 4 logs reduction in lentivector titre (Figures 2 y 3).

**Figure 2.**
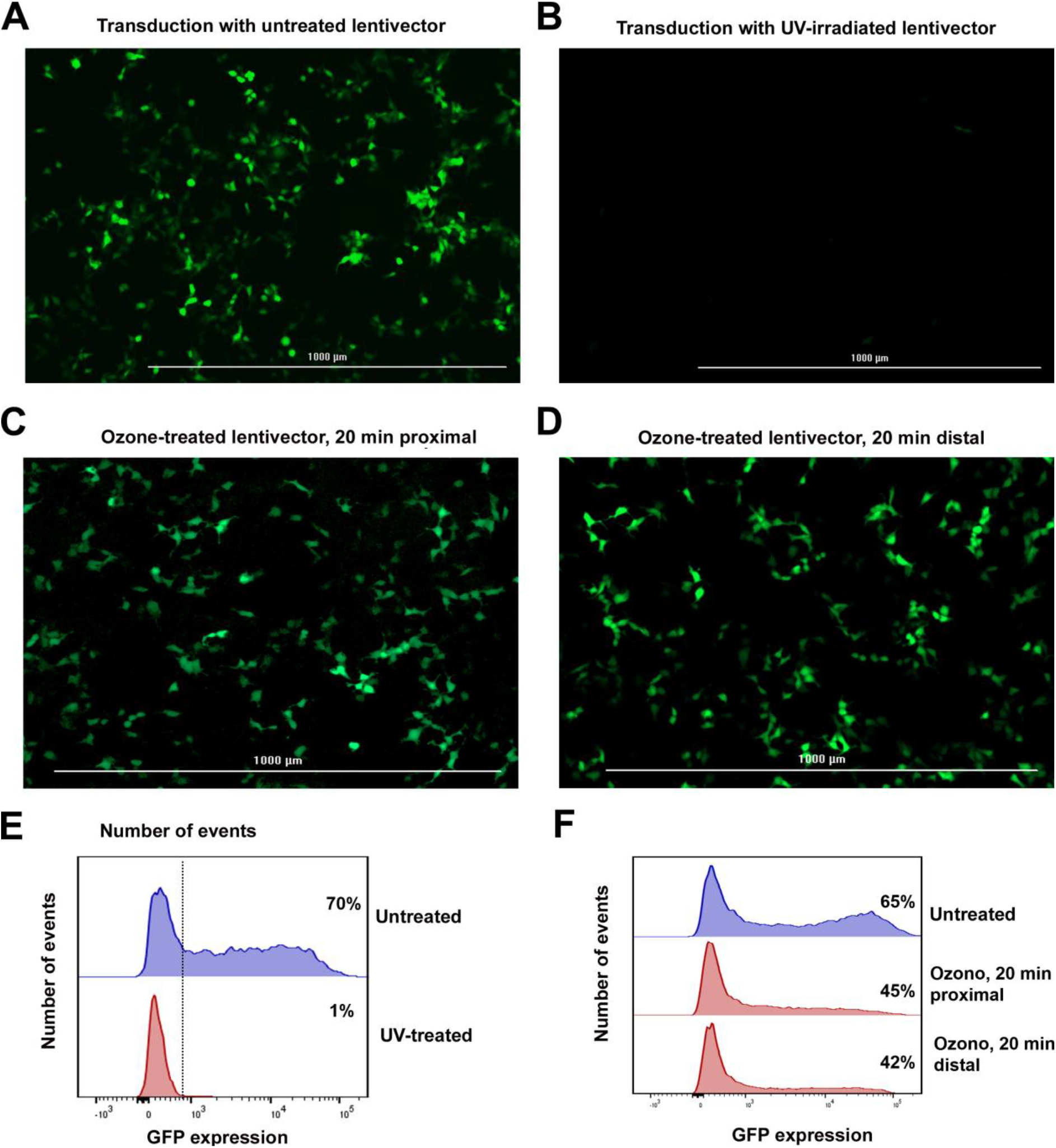
Effects of UV light and ozone treatments over the viability of lentivector preparations. ***(A)*** *Fluorescence microscopy picture of 293T cells transduced with an untreated control pSIN-GFP lentivector. GFP expression is indicative of infective capacities*. ***(B)*** *As in (A), but with lentivector stocks subjected to UV light treatments (UV-C, 20 min). The loss of viability is evident*. ***(C)*** *As in (B), but following a 20 min ozone treatment (>10 ppm) at the proximal location from the ozone cannon*. ***(D****) As in (C), but at the distal location*. ***(E)*** *Histograms with GFP expression from transduced 293T cells with the pSIN-GFP untreated lentivector stock, or previously treated with UV-light, as indicated. The vertical line indicates the separation between GFP positivity from negativity. Percentages of GFP-expressing 293T cells are shown within the graphs*. ***(F)*** *As in (E), but using lentivector stocks previously subjected to a 20 min ozone treatment at proximal and distal locations from the ozone cannon, as indicated*.

**Figure 3.**
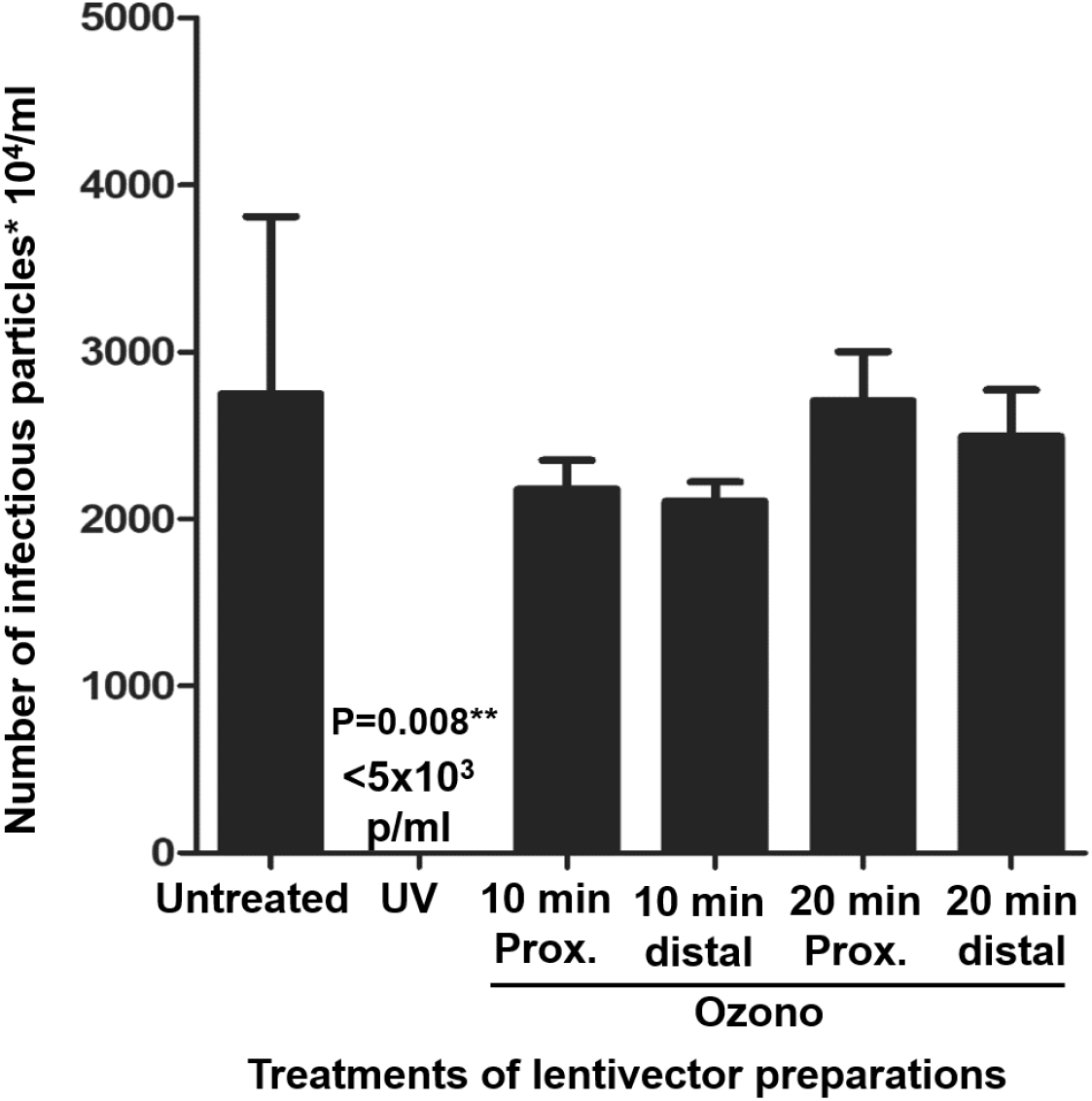
Titration of lentivector stocks after the treatments. *Bar graphs presenting the titres of the lentivector stocks after the indicated treatments. Only the UV-light treatment significantly decreased the viability of the lentivector preparations. The detection limit of the titration technique is shown within the graph (<5 × 10^3^ infectious particles/ml)*.

Importantly, no significant effects over the viability of bacterial samples were observed. The decrease in viable cells did not reach have a log for S. *aureus* MW2 and 1,4 logs for S. Enteritidis 3934. In contrast, UV irradiation reduced the number of viable cells below the detection limit of the titration technique (<10 Infectious colony-forming units /ml), reaching a decrease of at least 6 logs (Figure 4).

**Figure 4.**
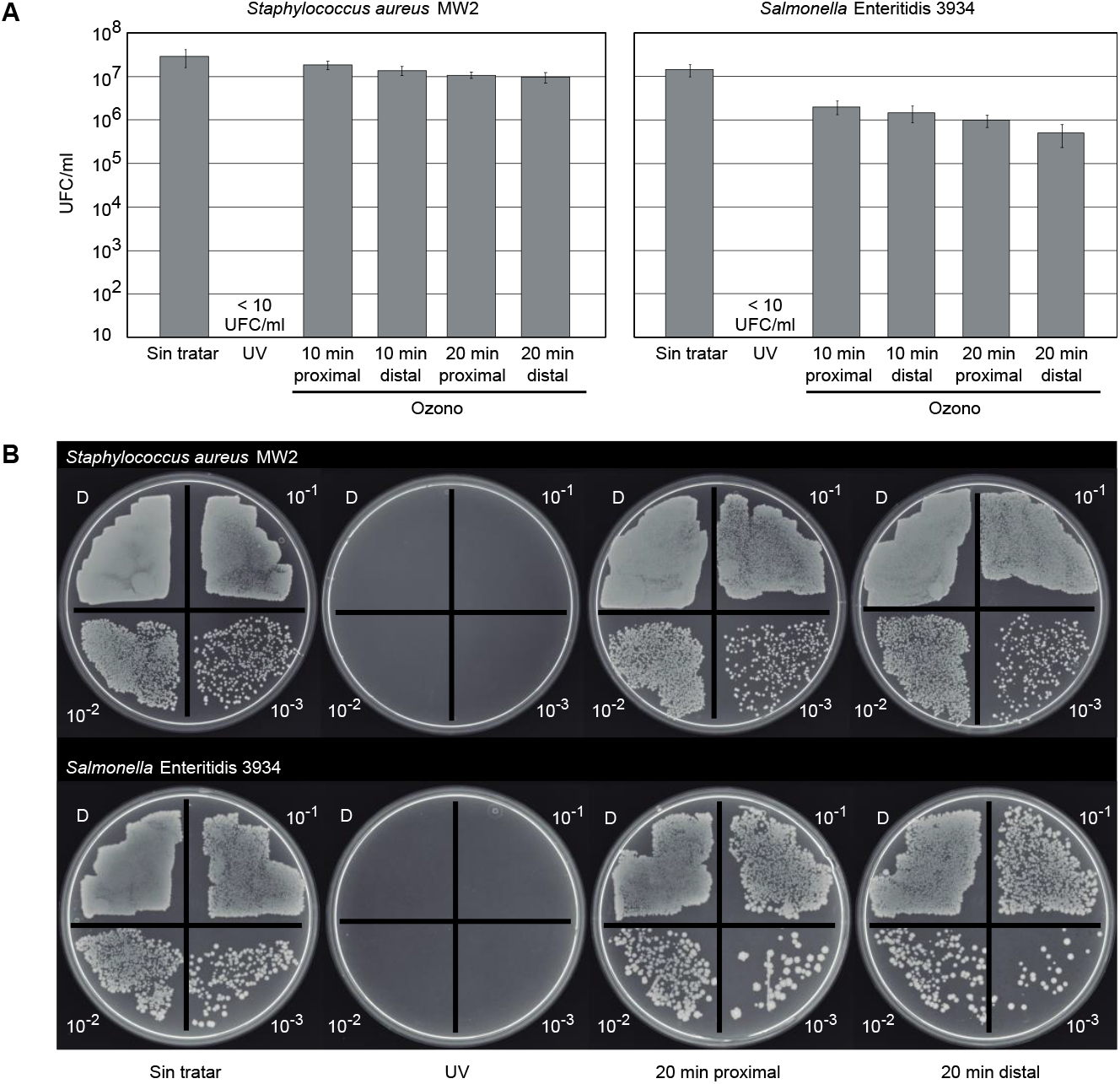
Effects of ozone treatments over the viability of S. aureus y S. Enteritidis. ***(A)*** *Infectious colony-forming units per ml of viable S. aureus MW2 y S. Enteritidis 3934 cells following the indicated treatments. Means and standard deviation error bars are shown, from three independent experiments. The detection limit of the quantification technique is shown (10 infectious colony-forming units/ml)*. ***(B)*** *Pictures of the culture dishes in which quantification of viable bacteria was peformed, by 10-fold serial dilutions for each bacterial strain. The dilution of the bacterial preparation seeded in each quandrant is indicated. “D” indicates the undiluted bacterial suspensión. Only the UV-light treatment significantly reduced the viability of both tested strains*.

These assays were repeated independently twice at different days, with equivalent results.

## DISCUSSION

Here we have used a lentivector preparation as a biosafe substitutive model for infectious SARS-CoV-2 virions to evaluate the disinfecting capacities of nebulized ozone. Lentivector particles share structural features that make them good candidates for infectious single-stranded RNA viruses such as coronaviruses. Lentivector preparations contain VLPs with packaged single-stranded RNA, protected with the nucleocapsid, capsid and matrix proteins, enveloped in a lipid membrane containing the VSV-G glycoprotein. In analogy, coronavirus particles are similar in size (100 nm diameter), with a single-stranded RNA genome packaged by the nucleoprotein. In turn, the viral nucleocapsid in enclosed within a lipid membrane in which the S, M and E proteins are incorporated. Due to the same chemical composition of their genomes (single-stranded RNA), we could hypothesize that both organisms as similarly susceptible to UV light and other oxidazing agents.

Guidelines of the Environmental Protection Agency (EPA, United States of America) (7) and the European Guidelines (EN-14885) reccomend a decrease superior than 3 logs and 5-6 logs for viruses and bacteria, respectively, for a disinfecting treatment to be considered efficacious by nebulization in hospitals. None of the two ozone treatments significantly reduced the lentivector titer (Figures 2 and 3), or S. *aureus* MW2 and S. Enteritidis 3934 bacterial counts (Figure 4). Therefore, these treatments cannot be considered effective for disinfecting contaminated surfaces in ambulances, in the tested conditions. Interestingly, the location of the samples from the ozone cannon did not influence the results, without significant reduction in lentivector treatments or bacterial counts (Figures 3 y 4).

The evidence from our study shows that gaseous ozone treatments currently applied in emergency vehicles do not significantly affect virus or bacteria viability. It has been previously shown virucide activities of gaseous ozone in surfaces at concentrations higher than 20-25 ppm in the presence of relative humidity higher than 90% (17). In our present study, we have quantified ozone concentrations higher than10 ppm, although we cannot discard that superior concentrations may have been reached as the upper detection limit of the ozometer is 10 ppm. Relative humidity at the time of the experiments ranged from 37% to 48%, which may have influenced the efficacy of the treatment. Nevertheless, it has to be remarked that these humidity levels correspond to those normally found within the vehicle, without using humidifyiers. The ozone is rapidly decomposed into highly oxidative OH radicals in the presence of water, while it is more stable in air. This difference in stability could explain the differences in efficacy found in water compared to nebulization.

Although we cannot discard that prolonged treatments with higher ozone concentrations may show some disinfecting capacities, our data indicates that the current treatments in emergency vehicles are insufficient. Therefore, we cannot recommend their use for this end.

## Data Availability

All data is available

## ACKNOWLEDGEMENTS

We would like to acknowledge the use of the test ambulance from Escuela Sanitaria Técnico Profesional de Navarra (ESTNA), and to Protección Civil de Navarra for the use of the ozone cannon.

## Conflict of interests

The authors disclose no conflicts of interest

## Notes

### Competing Interest Statement

The authors have declared no competing interest.

### Funding Statement

No external funding was received

### Author Declarations

This research is exempted from ethics committee

## REFERENCES

1. Gray NF. Chapter Thirty-Three—Ozone Disinfection. Second Edition. Microbiology of Waterborne Diseases: Microbiological Aspects and Risks. Elsevier; 2013. 17 p.

2. Ding W, Jin W, Cao S, Zhou X, Wang C, Jiang Q, et al. Ozone disinfection of chlorine-resistant bacteria in drinking water. Water Res. 2019 Sep 1;160:339–49.

3. Kim JG, Yousef AE, Dave S. Application of ozone for enhancing the microbiological safety and quality of foods: a review. J Food Prot. 1999 Sep;62(9):1071–87.

4. Naito S, Takahara H. Ozone Contribution in Food Industry in Japan. Ozone: Science & Engineering. 2006 Dec;28(6):425–9.

5. Escors D, Ortego J, Laude H, Enjuanes L. The Membrane M Protein Carboxy Terminus Binds to Transmissible Gastroenteritis Coronavirus Core and Contributes to Core Stability. J Virol. 2001 Feb 1;75(3):1312–24.

6. Escors D, Breckpot K. Lentiviral Vectors in Gene Therapy: Their Current Status and Future Potential. Arch Immunol Ther Exp (Warsz). 2010 Feb 9;58(2):107—19.

7. Green T. Product Performance Test Guidelines. 2018 Feb 20;:1–19.

8. Braunschweiler H. Technical Guidance Document in Support of the Directive 98/8/Ec Concerning the Placing of Biocidal Products on the Market Guidance on Data Requirements for Active Substances and Biocidal Products. 2008 Feb 1;:1–149.

9. Escors D, Lopes L, Lin R, Hiscott J, Akira S, Davis RJ, et al. Targeting dendritic cell signaling to regulate the response to immunization. Blood. 2008 Mar 15;111(6):3050–61.

10. Selden C, Mellor N, Rees M, Laurson J, Kirwan M, Escors D, et al. Growth factors improve gene expression after lentiviral transduction in human adult and fetal hepatocytes. J Gene Med. 2007 Feb;9(2):67–76.

11. Baba T, Takeuchi F, Kuroda M, Yuzawa H, Aoki K-I, Oguchi A, et al. Genome and virulence determinants of high virulence community-acquired MRSA. Lancet. 2002 May 25;359(9320):1819–27.

12. Solano C, García B, Valle J, Berasain C, Ghigo J-M, Gamazo C, et al. Genetic analysis of Salmonella enteritidis biofilm formation: critical role of cellulose. Mol Microbiol. 2002 Feb 1;43(3):793–808.

13. Rutala WA, Gergen MF, Weber DJ. Room Decontamination with UV Radiation. Infect Control Hosp Epidemiol. 2015 Jan 2;31(10):1025–9.

14. Weber DJ, Kanamori H, Rutala WA. “No touch” technologies for environmental decontamination. Current Opinion in Infectious Diseases. 2016 Aug;29(4):424–31.

15. Simmons S, Dale C, Holt J, Velasquez K, Stibich M. Role of Ultraviolet Disinfection in the Prevention of Surgical Site Infections. In: Ultraviolet Light in Human Health, Diseases and Environment. Cham: Springer International Publishing; 2017. pp. 255–66. (Advances in Experimental Medicine and Biology; vol. 996).

16. Boyce JM, Donskey CJ. Understanding ultraviolet light surface decontamination in hospital rooms: A primer. Infect Control Hosp Epidemiol. 2019 Jun 18;40(9):1030–5.

17. Hudson JB, Sharma M, Vimalanathan S. Development of a Practical Method for Using Ozone Gas as a Virus Decontaminating Agent. Ozone: Science & Engineering. 2009 May 29;31(3):216–23.

